# Cardiac arrest outcomes after targeted temperature management with an esophageal cooling device

**DOI:** 10.1101/19001487

**Authors:** Cedar Morrow Anderson, Rick Fisher, Donald Berry, J Brad Diestelhorst, Marvin Wayne

## Abstract

**Objective:** To assess the efficacy of an esophageal device to provide TTM (Target Temperature Management) post Cardiac Arrest

**Design:** A chart review of all patients treated with ETTM, following cardiac arrest. Initial patient temperature, time to target, supplemental methods (water blankets, head wraps, or ice packs), and patient survival were extracted for analysis.

**Setting:** Community Medical Center Intensive Care Unit

**Patients:** All patients receiving TTM via an esophageal device post Cardiac Arrest from August 2016 to November 2018

**Interventions:** TTM both cooling and warming via an esophageal device

**Measurements and Results:** A total of 54 patients were treated from August 2016 to November 2018; 30 received ETM only, 22 received supplemental cooling, and 2 had treatment discontinued prior to reaching target due to recovery. Target temperatures ranged from 32 to 36 degrees. The median time to target temperature for the entire cohort was 219 minutes (IQR 81-415). For the cohorts without, and with, supplemental cooling modalities, the median time to attain target temperature was 128 minutes (IQR 71-334), and 285 minutes (IQR 204-660), respectively. Survival to ICU discharge was 51.9% for the entire cohort.

**Conclusions:** ETM attains target temperature at a rate consistent with current guidelines and with similar performance to alternative modalities. This may provide a more cost-effective and approachable core cooling option to community hospitals that only use water blankets or other surface methods.

## INTRODUCTION

Targeted temperature management (TTM) is a long-standing standard of care for post-cardiac arrest patients.[1] However, consistently implementing a TTM protocol is challenging, especially in a community hospital. Often, the protocols described in the literature include labor- and cost-intensive methods, that are not feasible or sustainable in highly resource-constrained healthcare settings. For example, the intravascular cooling methods often outperform surface methods in terms of time to target temperature.[2-8] However, because intravascular cooling is physician initiated and introduces risks including catheter associated bloodstream infection (CLABSI), deep venous thrombosis (DVT) and pulmonary embolism (PE) [9-13], it may not be an ideal choice for some organizations. Surface devices can be deployed by a wider range of clinicians, such as nurses, and avoid CLABSI and reduce the risks of DVT and PE, but increase the incidence of shivering, which in turn impact temperature maintenance and adds to the overall cost of care.[14, 15]

Esophageal temperature management (ETM), a relatively new TTM method, offers minimally invasive core access. ETM uses a device that is placed in the esophagus, in a similar fashion to an orogastric tube. The device connects to external water blanket heat exchanger, which provides cold or warm water in a closed circuit, thereby transferring heat across the patient’s core. This approach avoids the risks of sterile intravascular placement and the challenges of transferring heat across the skin. Since our facility already owned compatible heat exchangers and these heat exchangers can be used simultaneously with our current stock of water blankets, the Value Analysis Committee found ETM to be a cost-effective option and approved implementation. The purpose of this study is to evaluate ETM performance, used alone or combined with surface methods, in a cohort of patients who received TTM after cardiac arrest.

## METHODS

### Study design

This was an IRB-approved (PHIRB 1085850-3) retrospective study of all patients treated with an ETM device (EnsoETM ECD02A, Attune Medical, Chicago, IL) for TTM after cardiac arrest in a single institution. All patients had attained return of spontaneous circulation (ROSC) after cardiac arrest from any rhythm, and were cooled according to local hospital protocol, using a Cincinnati Sub-Zero Blanketrol III Hyper-Hypothermia System connected to the ETM device. Additional surface wraps were added in patients at the discretion of the treating clinicians.

### Data collection

Charts were identified through a quality-control log of patients treated with temperature management. Details of treatment, including device used, temperature targeted, and time to target temperature were extracted by one study author (RF) and reviewed in cases of uncertainty by the remaining authors. Patient starting temperature, target temperature, and times of start and attainment of goal temperature were collected.

## STATISTICAL ANALYSIS

Data are reported with descriptive statistics, utilizing parametric or non-parametric measures where appropriate.

## RESULTS

A total of 54 patients were treated with ETM over the evaluation period from August 2016 to November 2018. Of these 54 patients, 30 had ETM only, 22 had supplemental, and 2 recovered and had treatment discontinued prior to reaching target. Target temperatures ranged from 32°C to 36°C, depending on clinician preference, with a range typically specified spanning 2-3 degrees (for example, 34−36°C, or 33°C−35°C).

The median time to attain target temperature for the entire cohort was 219 minutes (IQR 81-415). Patients treated with ETM only had a median time to target of 128 minutes (IQR 71-334) and patients who received with supplemental surface cooling had a median time to target temperature of 285 minutes (IQR 204-660). Survival to ICU discharge was 51.9% for the entire cohort, and survivors exhibited longer times to achieve goal temperature (median 180 minutes in non-survivors vs. 255 minutes in survivors).

## DISCUSSION

To the authors’ knowledge, this is the largest study to date characterizing time to target using ETM after cardiac arrest. The aim of the study was to determine cooling rate in a large cohort of patients treated at a single institution. Although the study was not designed to make a direct comparison between esophageal cooling and other modalities, the data suggest that the cooling rate is comparable to other methods. A review of independent published data on alternative cooling methods used in post-cardiac arrest TTM shows median times to target temperature ranging from 130 minutes to well over 600 minutes (Table I).

Some patients arrived at lower core temperatures than others, allowing them to reach target more quickly, while others took much longer to cool and required supplemental cooling via head wraps, chest wraps, blankets, or ice packs to achieve target temperature. While this retrospective study cannot identify the specific reasons that clinicians utilized additional cooling modalities for particular patients, our study found that survivors exhibited longer times to achieve goal temperature, a finding similar to other reports in which a quicker time to target temperature is associated with worse outcome.[18-21] In a study of 203 patients, non-survivors generated less heat than survivors and reached target temperature sooner (median, 2.2 hr [interquartile range, 1.5–3.8 hr] vs median, 3.6 hr [interquartile range, 2.0–5.0 hr]; p = 0.01).[18] A study of 177 patients found that median time to target temperature was shorter among non-survivors (200 [IQR 25–363] min) than survivors (270 [IQR 158–375] min, p = 0.03).[21] A study of 321 patients found that time to target temperature in the “good outcome” (CPC 1– 2) cohort was 237 (IQR142–361) minutes compared to 180 (IQR 100–276) minutes in the “poor outcome” (CPC 3–5) cohort (p = 0.004).[20] A study of 588 patients found that time to target temperature was 209 minutes (IQR 130-302) in patients with favorable neurological outcomes compared to 158 min (IQR 101-230) (P < 0.01) in patients with unfavorable neurological outcomes.[22] The hypothesis for these differences has been that those who cool more quickly have lost the ability to thermoregulate, which may represent more severe brain injury, and thus, worse outcomes, in this subset of patients.[18]

## LIMITATIONS

This study is limited by its retrospective nature; however, we utilized endpoints recorded in the medical record that are generally reliable and unlikely to be systematically biased. Although an inverse association was found between survival and cooling rate, no causation can be attributed.

## CONCLUSION

ETM attains target temperature with similar performance to alternative modalities, offering core patient cooling without the risks of intravascular placement. As in other studies, surviving patients required longer times to reach target temperature.

## Data Availability

All data used was de-identified and extracted with IRB permission from hospital records. We hold securely that de-identified data.

